# Genomic evolution and rearrangement of CTX-Φ prophage elements in Vibrio cholerae during the 2018-2022 cholera outbreaks in The Democratic Republic of Congo

**DOI:** 10.1101/2024.03.30.24305082

**Authors:** Leonid M. Irenge, Jérôme Ambroise, Bertrand Bearzatto, Jean-François Durant, Louisette K. Wimba, Jean-Luc Gala

## Abstract

Between 2018 and 2022, we conducted whole-genome sequencing and phylogenomic analysis on 173 *Vibrio cholerae* bacteria (172 *V. cholerae* O1 and 1 *V. cholerae* non-O1/non-O139 isolates) from cholera patients across four provinces in the Democratic Republic of Congo (North-Kivu, South-Kivu, Tanganyika, and Kasai Oriental). All *V. cholerae* O1 isolates were classified into the AFR10d and AFR10e sublineages of AFR10 lineage, originating from the third wave of the seventh El Tor cholera pandemic (7PET). Compared to the strains analysed between 2014 and 2017, we observed only limited genetic changes in the core genome of both sublineages. However, AFR10e expanded across the four provinces, whereas AFR10d appeared to become extinct by the end of 2020. The *V. cholerae* isolates from 2022 exhibited significant rearrangement in the CTX prophage and associated phage satellites. Notably, this included the tandem repeat of a novel environmental satellite phage RS1 downstream the *ctxB t*oxin gene of the CTX-Φ-3 prophage on the large chromosome, and two tandemly arrayed copies of the pre-CTX-Φ prophage precursor on the small chromosome. In conclusion, while the core genome of *V. cholerae* O1 AFR10d and AFR10e sublineages showed minimal changes, significant alterations in the content and organisation of elements associated with the CTX-Φ and pre-CTX prophages were identified in AFR10e *V. cholerae* O1 isolated in 2022.

## Introduction

Cholera, a severe watery diarrheal disease caused by *Vibrio cholerae* serogroups O1 and/or O139 (1, 2) remains a significant healthcare concern in developing countries (1, 3, 4). It is estimated to cause up to 3 million cases of cholera and approximately 95,000 deaths annually (5, 6). Over the past decade, Sub-Saharan Africa has become the most afflicted by cholera (7, 8), now referred to as the “new cholera homeland” (9), with countries in this region reporting 83% of global cases between 2000 and 2015 (10). The DRC is among the leading countries in terms of cholera cases, contributing 5-14% of the global case count annually (5, 11, 12). In 2020, DRC reported 19,789 cases, ranking second only to Yemen in terms of numbers (13). Recent data from 2023 indicate a deepening cholera crisis in the country, with over 41,000 cases and 314 deaths reported (14).

Cholera has traditionally been most prevalent in the Great Lakes Region (GLR) (16), but recent reports show it has spread to most DRC provinces, including areas well beyond the GLR (17). For example, between 2018-2022, Kasai Oriental province in the central region of DRC reported 2,176 cholera cases (18). The initial introduction of *V. cholerae* O1 strains into the DRC occurred during the first wave of P7ET, known as the T5 introduction (1970–1972) (19, 20). A subsequent introduction, in the early nineties, part of the P7ET’s third wave and designated as the T10 introduction event, brought a strain from southern Asia (21), which has persisted in the GRL through successive outbreaks (22, 23). Previous studies, including our own, have documented the division of the T10 (AFR10) lineage into two sublineages, AFR10d, and AFR10e, and identified a regional focus of endemic disease derived from the T10 introduction (22).

In this study, we conducted whole-genome sequencing and phylogenetic analyses on *V. cholerae* isolates collected from cholera cases in the North-Kivu, South-Kivu, and Tanganyika between 2018 and 2022, as well as from cases in Kasai Oriental specifically observed in 2020. By comparing these genomes with data from *V. cholerae* isolates collected between 2014 and 2017, we aim to assess the genetic evolution of *V. cholerae* in the DRC, and explore potential genetic links between isolates from the GLR and the central Kasai Oriental province.

## Material and methods

### Ethical clearance

The study protocols received approval from the Ethical Review Board (ERB) of the ISTM/Bukavu, DRC (ISTM-BUKAVU/CRPS/CIES/ML/0016/2023). The ERB granted a waiver of informed consent, determining that the study complied with international guidelines applicable to research during severe outbreaks. To protect confidentiality, samples were analysed anonymously.

### Phenotypic analysis

The isolation of *V. cholerae* bacteria and subsequent phenotypical tests and antimicrobial susceptibility assays followed protocols previously described (22). Rectal swabs were collected from patients exhibiting clinical symptoms of cholera, notably severe watery diarrhoea. These samples were incubated in alkaline peptone water for 6-8 hours, then streaked onto thiosulfate-citrate-bile salts-sucrose (TCBS) agar Petri dishes and incubated at 37°C overnight. Colonies appearing large yellow, flattened with opaque centres and translucent peripheries were further cultured on nutritive alkaline agar.

For phenotypical confirmation of *V. cholerae* standard microbiological tests were carried out (24), alongside immunological tests to determine the serotypes of the isolates. Antimicrobial susceptibility was assessed using the diffusion method on Mueller-Hinton agar plates, using antibiotic disks. Bacterial isolates were prepared at a density equivalent to 0.5 to 0.6 McFarland standard, and the zone of inhibition around antibiotics discs were measured after 16–20□hours of incubation.

Minimum inhibitory concentration (MIC) for ciprofloxacin was determined with MIC test strips (Liofilchem, Roseto degli Abruzzi, Italy) following the manufacturer’s guidelines. The interpretation of MIC values inhibition zone diameters was based on the EUCAST clinical breakpoint values (version 13.0, https://www.eucast.org/clinical_breakpoints).

### Whole-genome sequencing

Genomic DNA (gDNA) was isolated from all *V. cholerae* isolates in our collection using an EZ1 Advanced XL Biorobot and the Tissue DNA Kit (QIAGEN, Hilden, Germany), employing the Bacterial Card according as per the manufacturer’s instructions. Quantification of the isolated DNA was performed using a Qubit® fluorometer (Thermo Fisher Scientific, Eugene, OR, USA).

For whole-genome sequencing (WGS) of *V. cholerae,* we initially used Illumina short-reads sequencing. Short-read libraries were prepared from 70 ng DNA using an Illumina DNA Prep kit (Illumina, San Diego, CA, USA), and then sequenced using paired-end (2 x 300 bp) reads on a MiSeq platform (Illumina, San Diego, CA).

To investigate the genomic organisation within the prophage *V. cholerae* CTX-Φ, WGS was also carried out using the long-reads sequencing on the GridION platform (Oxford Nanopore Technologies, UK) for a subsect of 20 samples, selected across different collection years. Long-reads libraries were prepared from 400 ng of genomic DNA using the rapid barcoding kit 96 V14 (SQK-RBK114) and sequenced on an R10.4.1 flow cell run in a GridION device over a 72-hour period.

### Bioinformatics

Raw sequencing data from all *V. cholerae* isolates were submitted to the European Nucleotide Archive (ENA) under accession number PRJEB66194 (http://www.ebi.ac.uk/ena).

For Illumina short-reads, our bioinformatics pipeline started with quality checking via FastQC v.0.11.9 (25) followed by trimmomatic v.0.39 (26). De novo assembly of paired-end reads for each *V. cholerae* isolate was performed using Spades v.3.13.0 (27), generating a draft genome sequence. Assembly quality was assessed using QUAST 5.0.2 (28) and completeness was verified with the BUSCO v.4.1.1 algorithm, using the bacteria_odb10 dataset (29).

SNP-based phylogenetic analysis of *V. cholerae* genomes, including those from our previous (22) and other recent studies, was conducted using kSNP3.0, using the default settings, except for a k-mer size of 19 and the ‘-core’ option. Within identified clusters, representative genomes were selected, and a simplified phylogenomic tree was built using the R package ggtree (31). In-silico Multi-Locus Sequence Typing (MLST) was performed on all genomes by using the screen-Blast-mlst function of the custom Pathogenomics R package (https://github.com/UCL-CTMA/Pathogenomics), requiring 100% sequence identity and coverage for allele type assignment. All genomes were screened for virulence factor genes as per the VFDB database ((http://www.mgc.ac.cn/VFs) (32), with an identity cut-off set at 80%. Genotypic AMR profiles were assessed using AMRFinderPlus v3.10.24 and ARIBA v2.14.6.

Genomic organisation of the CTX-Φ prophage I, *V. Cholerae* O1 was determined using de novo assembled genomes from GridION long reads with canu v.2.2 (32). Representative sequences of the CTX-Φ prophage were localised using Blastn v.2.12.0 (33) in these contiguous genomes. Finally, the graphical representation of prophage organisation was created using the Gviz v.1.21.1 package in Bioconductor.

## Results

### V. cholerae serotyping and antimicrobial susceptibility

In this study, 173 clinical isolates of *V. cholerae* were evaluated. The phenotypic characterisation data and antimicrobial susceptibility profiles are presented in Table 1 (Table near here1).

**Table 1.** Antimicrobial susceptibility patterns and antimicrobial resistance (AMR) genes of *V. cholerae* isolates from eastern DRC. AMP: Ampicillin; CHL: Chloramphenicol; CIP: Ciprofloxacin; DOX: Doxycycline; ERY: Erythromycin; PXB: Polymyxin B; SXT: Sulfamethoxazole-Trimethoprim; TET: Tetracycline. Antimicrobial susceptibility profile of isolates is expressed as the percentage of resistant isolates of *V. cholerae*. AMR genes are in in italic.

| North-Kivu province 2018 (n = 19) |  |  |  |  |  |  |  |  |  |  |  |  |  |  |  |
| --- | --- | --- | --- | --- | --- | --- | --- | --- | --- | --- | --- | --- | --- | --- | --- |
| Serotype | Antimicrobial agents |  |  |  |  |  |  |  | AMR genes |  |  |  |  |  |  |
|  | AMP | CHL | CIP | DOX | ERY | PXB | SXT | TET | *gyrA* (S83I) | *parC* (S85L) | *SXT/R39I* | *catB9* | *floR* | *SulI* | *VarG* |
| O1 Inaba (n = 14) | S (100%) | S (100%) | S (100%) | S (100%) | S (100%) | R (100%) | R (100%) | S (100%) | 100% | 0% | 100% | 100% | 100% | 100% | 100% |
| O1 Ogawa (n = 5) | S (100%) | S (100%) | R (100%) | S (100%) | S (100%) | R (100%) | R (100%) | S (100%) | 100% | 100% | 100% | 100% | 100% | 100% | 100% |
| O1 non-O1/non-O139 (n = 0) | / | / | / | / | / | / | / | / | / | / | / | / | / | / | / |
| North Kivu province 2019 (n = 37) |  |  |  |  |  |  |  |  |  |  |  |  |  |  |  |
| Serotype | Antimicrobial agents |  |  |  |  |  |  |  | AMR genes |  |  |  |  |  |  |
|  | AMP | CHL | CIP | DOX | ERY | PXB | SXT | TET | *gyrA* (S83I) | *parC* (S85L) | *SXT/R39I* | *catB9* | *floR* | *SulI* | *VarG* |
| O1 Inaba (n = 10) | S (100%) | S (100%) | S (100%) | S (100%) | S (100%) | R (100%) | R (100%) | S (100%) | 100% | 0% | 100% | 100% | 100% | 100% | 100% |
| O1 Ogawa (n = 27) | S (100%) | S (100%) | R (100%) | S (100%) | S (100%) | R (100%) | R (100%) | S (100%) | 100% | 100% | 100% | 100% | 100% | 100% | 100% |
| O1 non-O1/non-O139 (n = 0) | / | / | / | / | / | / | / | / | / | / | / | / | / | / | / |
| North-Kivu province 2020 (n = 16) |  |  |  |  |  |  |  |  |  |  |  |  |  |  |  |
| Serotype | Antimicrobial agents |  |  |  |  |  |  |  | AMR genes |  |  |  |  |  |  |
|  | AMP | CHL | CIP | DOX | ERY | PXB | SXT | TET | *gyrA* (S83I) | *parC* (S85L) | *SXT/R39I* | *catB9* | *floR* | *SulI* | *VarG* |
| O1 Inaba (n = 11) | S (100%) | S (100%) | S (100%) | S (100%) | S (100%) | R (100%) | R (100%) | S (100%) | 100% | 0% | 100% | 100% | 100% | 100% | 100% |
| O1 Ogawa (n = 15) | S (100%) | S (100%) | R (100%) | S (100%) | S (100%) | R (100%) | R (100%) | S (100%) | 100% | 100% | 100% | 100% | 100% | 100% | 100% |
| O1 non-O1/non-O139 (n = 0) | / | / | / | / | / | / | / | / | / | / | / | / | / | / | / |
| North-Kivu province 2022 (n = 26) |  |  |  |  |  |  |  |  |  |  |  |  |  |  |  |
| Serotype | Antimicrobial agents |  |  |  |  |  |  |  | AMR genes |  |  |  |  |  |  |
|  | AMP | CHL | CIP | DOX | ERY | PXB | SXT | TET | *gyrA* (S83I) | *parC* (S85L) | *SXT/R39I* | *catB9* | *floR* | *SulI* | *VarG* |
| O1 Inaba (n = 0) | / | / | / | / | / | / | / | / | / | / | / | / | / | / | / |
| O1 Ogawa (n = 26) | S (100%) | S (100%) | R (100%) | S (100%) | S (100%) | R (100%) | R (100%) | S (100%) | 100% | 100% | 100% | 100% | 100% | 100% | 100% |
| O1 non-O1/non-O139 (n = 0) | / | / | / | / | / | / | / | / | / | / | / | / | / | / | / |
| South-Kivu province 2018 (n = 11) |  |  |  |  |  |  |  |  |  |  |  |  |  |  |  |
| Serotype | Antimicrobial agents |  |  |  |  |  |  |  | AMR genes |  |  |  |  |  |  |
|  | AMP | CHL | CIP | DOX | ERY | PXB | SXT | TET | *gyrA* (S83I) | *parC* (S85L) | *SXT/R39I* | *catB9* | *floR* | *SulI* | *VarG* |
| O1 Inaba<br>(n = 2) | S (100%) | S (100%) | S (100%) | S (100%) | S (100%) | R (100%) | R (100%) | S (100%) | 100% | 0% | 100% | 100% | 100% | 100% | 100% |
| O1 Ogawa<br>(n = 9) | S (100%) | S (100%) | R (100%) | S (100%) | S (100%) | R (100%) | R (100%) | S (100%) | 100% | 100% | 100% | 100% | 100% | 100% | 100% |
| O1 non-O1/non-O139<br>(n = 0) | / | / | / | / | / | / | / | / | / | / | / | / | / | / | / |
| **South-Kivu province 2019 (n = 5)** |  |  |  |  |  |  |  |  |  |  |  |  |  |  |  |
| Serotype | Antimicrobial agents |  |  |  |  |  |  |  | AMR genes |  |  |  |  |  |  |
|  | AMP | CHL | CIP | DOX | ERY | PXB | SXT | TET | *gyrA*<br>(S83I) | *parC*<br>(S85L) | SXT/R39I | *catB9* | *floR* | *SulI* | *VarG* |
| O1 Inaba<br>(n = 0) | / | / | / | / | / | / | / | / | / | / | / | / | / | / | / |
| O1 Ogawa<br>(n = 26) | S (100%) | S (100%) | R (100%) | S (100%) | S (100%) | R (100%) | R (100%) | S (100%) | 100% | 100% | 100% | 100% | 100% | 100% | 100% |
| O1 non-O1/non-O139<br>(n = 0) | / | / | / | / | / | / | / | / | / | / | / | / | / | / | / |
| **South-Kivu province 2020 (n = 20)** |  |  |  |  |  |  |  |  |  |  |  |  |  |  |  |
| Serotype | Antimicrobial agents |  |  |  |  |  |  |  | AMR genes |  |  |  |  |  |  |
|  | AMP | CHL | CIP | DOX | ERY | PXB | SXT | TET | *gyrA*<br>(S83I) | *parC*<br>(S85L) | SXT/R39I | *catB9* | *floR* | *SulI* | *VarG* |
| O1 Inaba<br>(n = 0) | / | / | / | / | / | / | / | / | / | / | / | / | / | / | / |
| O1 Ogawa<br>(n = 19) | S (100%) | S (100%) | R (100%) | S (100%) | S (100%) | R (100%) | R (100%) | S (100%) | 100% | 100% | 100% | 100% | 100% | 100% | 100% |
| O1 non-O1/non-O139<br>(n = 1) | S | S | S | S | S | S | S | S | / | / | / | / | / | / | / |
| **South-Kivu province 2022 (n = 27)** |  |  |  |  |  |  |  |  |  |  |  |  |  |  |  |
|  | Antimicrobial agents |  |  |  |  |  |  |  | AMR genes |  |  |  |  |  |  |
|  | AMP | CHL | CIP | DOX | ERY | PXB | SXT | TET | *gyrA*<br>(S83I) | *parC*<br>(S85L) | SXT/R39I | *catB9* | *floR* | *SulI* | *VarG* |
| O1 Inaba<br>(n = 0) | / | / | / | / | / | / | / | / | / | / | / | / | / | / | / |
| O1 Ogawa<br>(n = 27) | S (100%) | S (100%) | R (100%) | S (100%) | S (100%) | R (100%) | R (100%) | S (100%) | 100% | 100% | 100% | 100% | 100% | 100% | 100% |
| O1 non-O1/non-O139<br>(n = 0) | / | / | / | / | / | / | / | / | / | / | / | / | / | / | / |
| **Kasai-Oriental province 2020 (n = 9)** |  |  |  |  |  |  |  |  |  |  |  |  |  |  |  |
| Serotype | Antimicrobial agents |  |  |  |  |  |  |  | AMR genes |  |  |  |  |  |  |
|  | AMP | CHL | CIP | DOX | ERY | PXB | SXT | TET | *gyrA*<br>(S83I) | *parC*<br>(S85L) | SXT/R39I | *catB9* | *floR* | *SulI* | *VarG* |
| O1 Inaba<br>(n = 0) | / | / | / | / | / | / | / | / | / | / | / | / | / | / | / |
| O1 Ogawa<br>(n = 27) | S (100%) | S (100%) | R (100%) | S (100%) | S (100%) | R (100%) | R (100%) | S (100%) | 100% | 100% | 100% | 100% | 100% | 100% | 100% |
| O1 non-O1/non-O139<br>(n = 0) | / | / | / | / | / | / | / | / | / | / | / | / | / | / | / |
| **Tanganyika province 2022 (n = 3)** |  |  |  |  |  |  |  |  |  |  |  |  |  |  |  |

| Serotype | Antimicrobial agents |  |  |  |  |  |  |  | AMR genes |  |  |  |  |  |  |
| --- | --- | --- | --- | --- | --- | --- | --- | --- | --- | --- | --- | --- | --- | --- | --- |
| AMP | CHL | CIP | DOX | ERY | PXB | SXT | TET | *gyrA* (S83I) | *parC* (S85L) | SXT/R39I | *catB9* | *floR* | *SulI* | *VarG* | AMP |
| O1 Inaba<br>(n = 0) | / | / | / | / | / | / | / | / | / | / | / | / | / | / | / |
| O1 Ogawa<br>(n = 3) | S (100%) | S (100%) | R (100%) | S (100%) | S (100%) | R (100%) | R (100%) | S (100%) | 100% | 100% | 100% | 100% | 100% | 100% | 100% |
| O1 non-O1/non-O139<br>(n = 0) | / | / | / | / | / | / | / | / | / | / | / | / | / | / | / |

Of these isolates, 172 tested positive for agglutination with the O1 antiserum. Among these, 145 isolates agglutinated with the Ogawa and 27 with the Inaba antisera. All 172 *V. cholerae* isolates O1 were found to be susceptible to ampicillin, chloramphenicol, and tetracycline. However, they showed resistance to cotrimoxazole and polymyxin B, the latter being indicative of the El Tor biotype of *V. cholerae* (34). While all Inaba isolates (n = 27) were susceptible to ciprofloxacin, the Ogawa isolates (n = 146) demonstrated reduced susceptibility to this antibiotic. The non-O1/non-O139 isolate was susceptible to all antimicrobial agents tested including polymyxin B.

### Phylogenetics of V. cholerae O1 isolates in the DRC provinces under study

Phylogenetic analyses revealed that all 172 *V. cholerae* O1 isolates belonged to the AFR10 lineage (comprising AFR10d and AFR10e sublineages), aligning with previous reports from the same region (22, 23). Consistent with an earlier study, DRC AFR10 *V. cholerae* O1 isolates were divided into two sublineages: AFR10e and AFR10d, associated with sequence types ST69 and ST515, respectively as shown in Fig. 1 (insert figure 1 near here).

**Figure 1.**
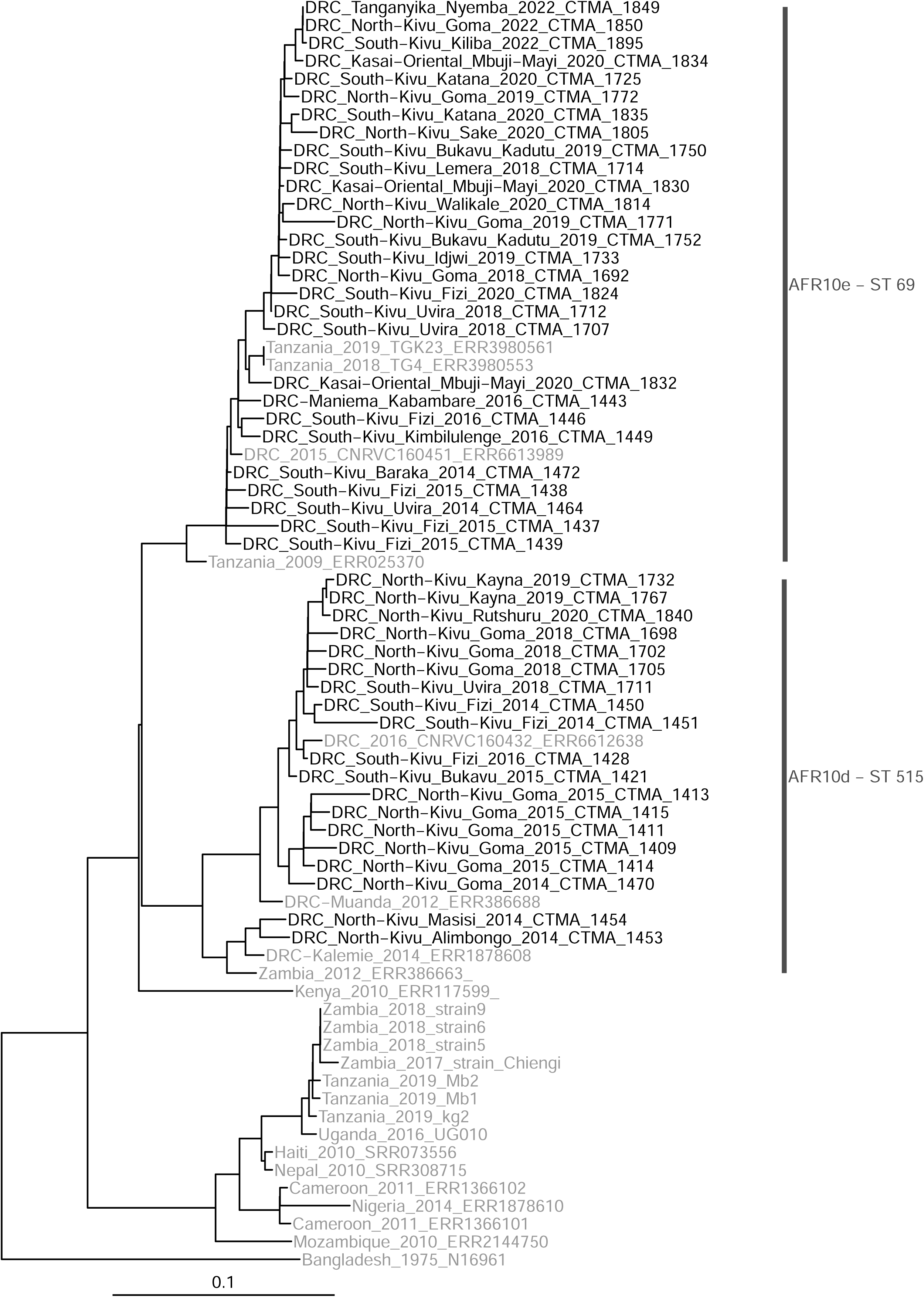
Phylogenomic analysis of *V. cholerae* O1 isolates (2014–2022) from four investigated DRC provinces. The 7PET *V. cholerae* O1 biotype EL Tor N19691 was used as an outgroup to root the tree. The scale bar represents substitutions per variable site in the core genome. *V. cholerae* O1 isolates from DRC are in black, while isolates from other African countries and the isolate N19691 are in grey.

All isolates from the Kasai Oriental (n = 9) and Tanganyika provinces (n = 2) characterised in this study belonged to the AFR10e sublineage (Fig.2) (insert figure 2 near here). From the 62 *V. cholerae* O1 isolates characterised in South-Kivu province between 2018 and 2022, the vast majority (n = 60) belonged to the AFR10e lineage, whereas only two isolates (collected in 2018) belonged to the AFR10d sublineage. Similarly, in North Kivu, the incidence of AFR10d declined dramatically by the end of 2019, with only one AFR10d isolate identified in 2020, and none reported since. During the 2018-2022 period, there were 73 isolates from the AFR10e and 25 from the AFR10d sublineage. Therefore, in contrast to our observations during the 2014-2017 period, the AFR10e sublineage significantly outpaced its AFR10d counterpart in both North-Kivu and South-Kivu provinces (see Fig. 2).

**Figure 2.**
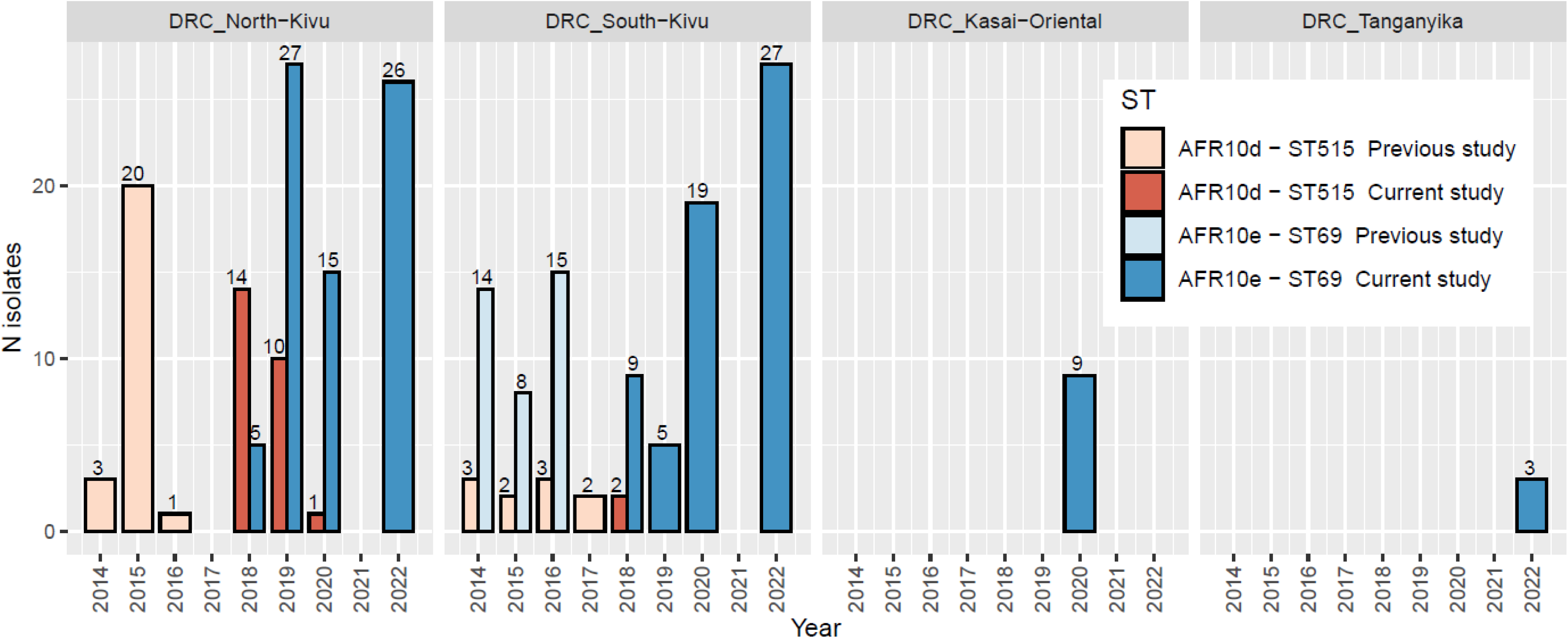
Temporal prevalence of AFR10d and AFR10e *V. cholerae* O1 isolates in the four DRC provinces under investigation. For the Kasai-Oriental and Tanganyika provinces, strains were collected only in 2020 and 2022, respectively.

### AMR genes

The analysis of antimicrobial (AMR) genes revealed that all DRC AFR10d and AFR10e isolates, consistent with previous studies, harboured the SXT/R391 integrative conjugative element ICEVchBan5, which carries genes associated with reduced susceptibility to cotrimoxazole (35) (Table 1). Similar to findings in all isolates of the AFR10 sublineage (23), both AFR10d and AFR10e isolates exhibited the S83I substitution in the gyrA protein.

Additionally, all AFR10e isolates presented the S85L substitution in the parC protein, a combination associated with reduced susceptibility to quinolones in *V. cholerae* (23). Notably, no AFR10d isolate in this study was found to carry both the S83I substitution in gyrA and the S85L substitution in parC. This observation was also true for clinical *V. cholerae* isolates from Tanganyika (n = 3), and Kasai Oriental (n = 9), all phylogenetically belonging to the AFR10e sublineage, and carrying both the S83I gyrA, and the S85L parC substitutions. Moreover, all *V. cholerae* O1 isolates characterised in this study possessed the *cat*B9, and *flo*R AMR genes, which confer resistance to chloramphenicol, the beta-lactamase *var*G gene, and the *sulI* gene associated with sulphonamide resistance. Surprisingly, the presence of these AMR genes did not compromise the susceptibility of AFR10 isolates to ampicillin, chloramphenicol, and tetracycline.

### Virulence genes

AFR10d and AFR10e isolates analysed in DRC between 2018 and 2022 possessed the same virulence genes as those characterised in the earlier 2014-2017 study (22). These include:

1. Virulence genes associated with the CTX-Φ-3 prophage and its phage satellites (36, 37), as previously described in all AFR10 isolates (21);
2. The complete Vibrio pathogenicity island-1 (VPI-1) (38), the Vibrio Seventh Pandemic Island-I (VSP-I), as well as the Vibrio Seventh Pandemic Island (VSP-II) (39). Notably, VSP-II exhibits a deletion spanning from ORF VC_0495 to VC_0512.

Furthermore, no genetic changes were identified in the virulence genes compared to those in the previous study (22).

### CTX-**Φ** prophage organization in O1 isolates

Long reads sequencing of 20 out of 172 *V. cholerae* O1 isolates using the GridION ONT platform, followed by assembly with Canu, generated contiguous genomes for each isolate. This approach facilitated the comprehensive characterisation of repetitive elements within the CTX-Φ prophage.

Isolates collected between 2018 and 2020 (Fig. 3A) (insert figure 3A near here). harboured a complete CTX-Φ-3 prophage, which includes the RS2 satellite phage and the core CTX-Φ prophage carrying a classical *ctx*B gene. This prophage was integrated into the large chromosome between the loci VC1465 and the *rtx*A gene of the RTX toxin locus. In most of these isolates, the CTX-Φ-3 prophage was flanked upstream by one canonical RS1 satellite phage (RS1^ca^), aligning with observations of several CTX-Φ prophages in atypical El Tor *V. cholerae* strains (36, 37).

**Figure 3A.**
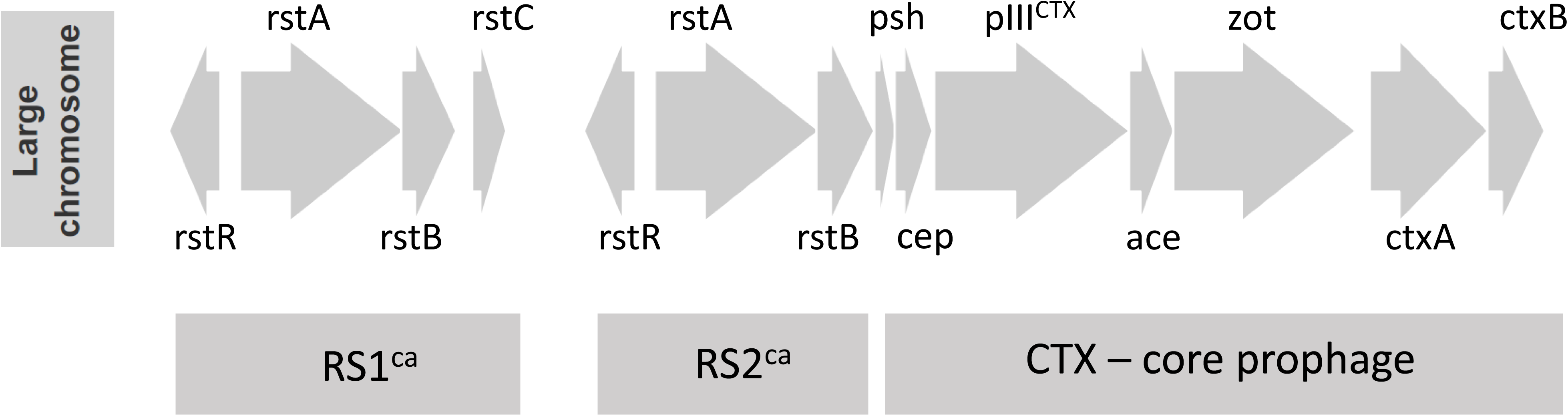
Representative organisation of the CTX-Φ prophage and its satellite in *V. cholerae* isolates collected between 2018 and 2020 in the DRC. Bloc arrows represent genes present in each element of the CTX-Φ-3-phage and indicate the direction of transcription. RS1^ca^ and RS2^ca^ represent RS1 and RS2 canonical satellite phages.

Isolates collected in 2022 displayed significant changes in the organisation of the CTX-Φ prophage array and its satellites on both the large and the small chromosomes. On the large chromosome, one RS1^ca^ satellite phage flanked the CTX-Φ-3 prophage upstream, while two tandem copies of an environmental RS1, named *RS1^env-DRC^*, flanked the CTX-3 Φ prophage downstream of the *ctx*B toxin gene (Fig. 3B) (insert figure 3B near here). The RS1^env-DRC^ variant is characterised by canonical *rst*A and *rst*B genes, an *rstC* gene with a single amino acid mismatch (L64P) compared to the canonical *rstC* gene in CTX-Φ prophage, and an *rstR* gene coding for a 86-aa long protein that matches perfectly (100%) with the rstR characterised in the environmental Mozambican *V. cholerae* IB1617 isolate (40).

**Figure 3B.**
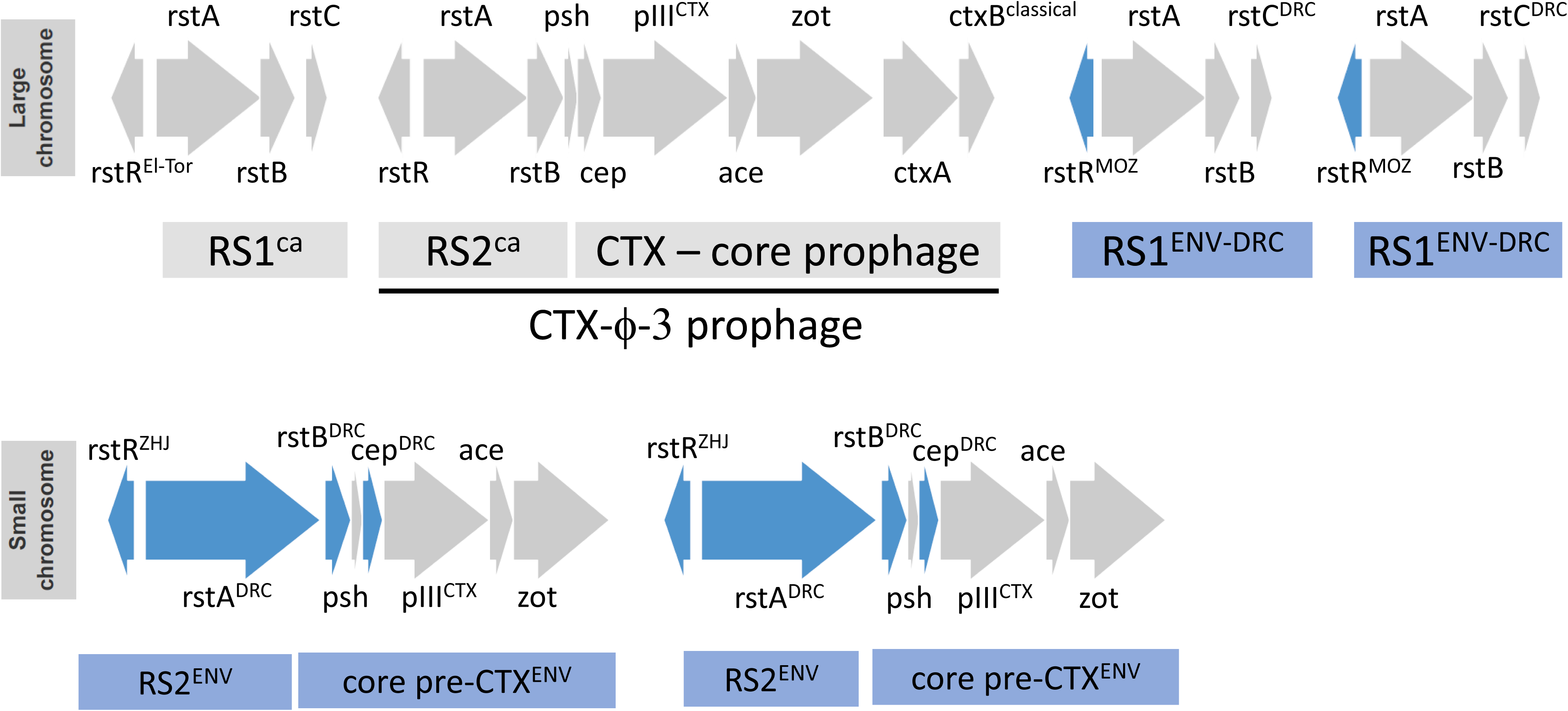
Representative organisation of the CTX-Φ prophage and its satellites in both the large and small chromosomes in *V. cholerae* isolates collected in 2022 in the DRC. Canonical sequences for each gene are shown in grey, while environmental sequences are shown in blue.

Another notable finding was observed in DRC *V. cholerae* O1 bacteria isolated from 2022, specifically on the small chromosome with the presence of two tandemly arrayed copies of a pre-CTX-Φ prophage inserted between VCA0569 and VCA0570 loci (37) (Fig. 3B). This arrangement resembles the configuration of the pre-CTX-Φ prophage on the small chromosome of the *V. cholerae* O139 VC1374 (41). Despite sharing a similar pre-CTX-Φ prophage configuration and an identical *rst*R gene (named *rst*R^ZHJ^) with the *V. cholerae* O139 VC1374 isolate, the 2022-DRC *V. cholerae* O1 isolates exhibited several amino acid substitutions in the rstA, rstB and cep proteins compared to both the *V. cholerae* O139 VC1374, and to *V. cholerae* non-O1/non-O139 VCE232 (42). These variants in the rstA, rstB, and cep proteins have been designated as rstA^DRC^, rstB^DRC^, and cep^DRC^, respectively, to distinguish them from their counterparts in the environment.

Consequently, *V. cholerae* O1 isolates collected in 2022 demonstrated a unique configuration of the CTX-Φ prophage array and its satellites on the large chromosome (RS1^ca^ - RS2^ca^ - *psh - cep - pIII^ctx^ - ace - zot - ctx*A - *ctx*B^classical^ - *RS1^env-DRC^*- *RS1^env-DRC^*), and on the small chromosome (a tandem arrangement of *rst*R^ZHJ^ - *rst*A^DRC^ *- rst*B^DRC^ - *psh - cep*^DRC^ - *pIII^ctx^ - ace - zot*). Notably, such a hybrid configuration has not been previously reported in 7PET *V. cholerae* O1 from the third wave.

### Characterization of the non-O1/non-O139 V. cholerae isolate

In this study, a single non-O1/non-O139 *V. cholerae* bacterium, isolated from a patient residing the city of Fizi, on the west bank of Lake Tanganyika, was characterized. This isolate exhibited a novel allele of the housekeeping *pyr*C gene, which led to designation as ST 362 by the curator of the MLST database. The combination of this allele with existing alleles for the other housekeeping genes constitutive of the MLST typing scheme (*adk* = 96, *gyr*B = 41; *mdh* = 109; *met*E = 237; *pnt*A = 136; *pur*M = 1*; pyr*C = 362) resulted in a new MLST profile, assigned the number 1577.

Similar to other non-O1/O139 *V. cholerae* isolates previously identified in the DRC (22), this ST1577 isolate lacked most PAI genes characteristic of the 7PET. Nonetheless, it harboured several virulence genes, including those related to Type VI Secretion System (T6SS) - VCA0109, VCA0122, vgrG.2 and *vgr*G.3 -, the *rtx* cluster - *rtx*A, *rtx*B, rtxC, *rtx*D genes -, and the virulence-associated (*vas*) operon.

## Discussion

The objective of this study was to phylogenetically characterise *V. cholerae* isolates associated with cholera outbreaks in DRC from 2018 to 2022, building upon a previous study that analysed isolates from the eastern DRC between 2014 and 2017 (22). Our results are consistent with previous reports that *V. cholerae* O1 isolates from the third wave and T10 transmission (AFR10) remain the major clade responsible for cholera outbreaks in the eastern part of the country (13, 22, 23). This evidence marks the sustained prevalence of a single clade as unprecedented when compared to the establishment of other V*. cholerae* O1 clades in neighbouring African countries (43–45).

Furthermore, our findings are in line with recent studies that show an AFR10d decline and the dominance of the AFR10e sublineage (23). In particular, we demonstrate that *V. cholerae* O1 isolates from Kasaï Oriental province, the first from central DRC to be molecularly characterised, belong to the AFR10e sublineage. These isolates are genetically linked to cholera outbreaks in the GLR provinces, highlighting the geographical spread and persistence of this clade within the DRC.

Despite geographic isolation of the Kasai Oriental province, which lacks direct natural linkage to the GLR through river or lake, the identification of AFR10e isolates in this area suggests potential transmission routes between the GLR and central regions. Human movement, including train travel, could facilitate such transmission. However, AFR10e *V. cholerae* bacteria isolated in 2022 exhibited significant genetic changes, in particular in the CTX-Φ prophage configuration. Although rearrangements in CTX-Φ prophage have often been linked to specific waves of *V. cholerae* O1 (36), our results show that all analysed *V. cholerae* O1 isolates still belong to the 10^th^ transmission event.

Noteworthy, an upstream RS1^ca^ satellite and two downstream tandem copies of RS1^env-DRC^, a novel environmental RS1 satellite, flanked the CTX-Φ-3 prophage on the large chromosome. Additionally, two tandemly arranged pre-CTX-Φ^env^ prophages containing the rstR^ZHJ^ sequence, similar to those described in a *V. cholerae* O139 isolate (41), were discovered on the small chromosome of these DRC *V. cholerae* O1 isolates. This rearrangement results in the co-existence of a CTX-3-Φ prophage, pre-CTX-Φ prophage, and RS1 satellites with both clinical and environmental origins within the same bacterial cell. Considering the crucial role of RS1 and pre-CTX-Φ proteins in regulating the replication, morphogenesis, and the virulence of the CTX-Φ phage (46), these observed genomic rearrangements may significantly influence the evolutionary dynamics of *V. cholerae*, including replication, excision, and the horizontal transfer of CTX-Φ prophage elements.

The discovery of these genetic changes in AFR10e isolates collected in 2022 from various provinces across the DRC not only underscores their expansion into new territories but also suggests an enhanced capacity for environmental adaptation. This adaptability may provide them with a competitive advantage over *V. cholerae* O1 isolates from previous years (2014–2021), potentially facilitating their rapid spread and dominance.

In conclusion, our study not only confirms the continued dominance of AFR10e sublineage in cholera outbreaks within the DRC but also highlights potential transmission routes and reveals significant genetic alterations within the CTX-Φ prophage. These findings underscore the critical need for ongoing monitoring of *V. cholerae* O1 to better predict and mitigate future outbreaks, emphasising the importance of understanding genetic evolution in the effectiveness of public health responses.

## Data Availability

All the data produced in the present work are contained in the manuscript

https://www.eucast.org/clinical_breakpoints

http://www.mgc.ac.cn/VFs

https://github.com/UCL-CTMA/Pathogenomics

## Acknowledgments

We extend our gratitude to the technical teams of C.D.R.M/AFIA-TELE (South-Kivu Province) and AMI-LABO (North-Kivu province) for their invaluable assistance in the sampling and processing of faecal samples from patients suspected of having cholera, which facilitated the isolation of *V. cholerae*. Special thanks are due to the laboratories and authors who submitted sequences to the GISAID EpiFlu and the NCBI databases.

Further acknowledgment goes to the EMBL-EBI team (Genome Campus, Hinxton, Cambridgeshire, CB10 1SD, UK) for providing ENA accession numbers, thus enhancing the accessibility of our data. We are particularly grateful to Dr. Sophie Octavia, the curator of the *Vibrio cholerae* MLST database, for her expertise and assistance in assigning the new MLST profile.

